# Automated Identification of Complex Percutaneous Coronary Intervention from Cardiac Catheterization Reports using Large Language Models

**DOI:** 10.64898/2026.08.05.26359802

**Authors:** Nilay Bhatt, Fred Warner, Jennifer Miao, Ravi Thakker, Golsa Joodi, Pablo Cantero-Schaffer, Bobak Mortazavi, Chenxi Huang, Harlan M Krumholz, Karthik Murugiah

## Abstract

**Introduction:** Manual abstraction of complex percutaneous coronary intervention (PCI) variables from cardiac catheterization reports is labor-intensive and limits scalable cardiovascular research. We evaluated open-weight large language models (LLMs) for automated complex PCI phenotyping.

**Methods:** We evaluated three LLMs (Llama 3.3 70B, Meditron-7B, and BioMistral-7B) using manually annotated catheterization reports from three hospitals within Yale New Haven Health. Models identified PCI reports and extracted six complex PCI features: 3 vessels treated, ≥3 lesions treated, bifurcation PCI with two stents, chronic total occlusion, ≥3 stents, and total stent length ≥60 mm.

**Results:** Among 1,412 clinical notes, 596 were PCI reports. Llama 3.3 70B outperformed the smaller domain-specific models across most tasks. For PCI identification, Llama 3.3 70B achieved 100.0% sensitivity, 93.8% specificity, 96.4% accuracy, and 95.9% F1 score. Among 590 evaluable PCI reports (excluding 6 indeterminable cases due to missing variables) for complex PCI classification, Llama 3.3 70B achieved 97.7% sensitivity, 80.1% specificity, 57.6% positive predictive value, 99.2% negative predictive value, 83.9% accuracy, and 72.5% F1 score. Performance was higher for explicitly documented variables, including stent number and length, and lower for variables requiring interpretation across procedural details, including lesion count, bifurcation PCI, and chronic total occlusion PCI. Llama 3.3 70B had the highest accuracy at each site for complex PCI classification but significant site-level heterogeneity was observed.

**Conclusion:** A high-capacity open-weight LLM accurately extracted complex PCI variables from unstructured reports and outperformed smaller domain-specific models. These findings support the potential use of locally deployable LLMs for scalable automated PCI phenotyping.

## INTRODUCTION

Cardiac catheterization and percutaneous coronary intervention (PCI) procedure notes contain rich data regarding coronary anatomy, procedural technique, device parameters, and angiographic outcomes. This information is invaluable for research, however, extracting this information is a resource intensive process^1^ due to the unstructured free-text format of PCI procedure notes. Registries like the National Cardiovascular Data Registry (NCDR) CathPCI registry^2^ systematically collect data related to the PCI to support both quality improvement and research. Submitting such data to registries, however, can be a high-cost investment for hospitals with specialized staff requirements.

The advent of large language models (LLMs) has made it possible to automate information extraction from complex free-text notes such as PCI procedure notes without significant programming effort to build custom algorithms etc.^3,4^ Further, the reasoning abilities of LLMs open the possibility of collecting nuanced data which could previously only be obtained via human review and interpretation.^4^ LLMs also reduce the reliance on consistent templates which make these algorithms transportable across institutions and ideal for at-scale information extraction.^5^ However, there is little data on how LLMs perform in extracting information from cardiac catheterization notes, especially complex variables.^1,6^

We evaluate the performance of state-of-the-art LLMs to extract information from PCI procedure notes in the use-case of ‘complex PCI’. Complex PCI as defined by Giustino et. al.^7^ is a clinically meaningful procedural phenotype associated with increased technical difficulty, adverse events, and resource utilization. Operational definitions of complex PCI are frequently used in outcomes research and quality programs to stratify risk and characterize case mix. The six component features of complex PCI are recorded in free-text PCI procedure notes and require a degree of domain expertise for a human annotator to extract.

## METHODS

A schematic overview of the study process is shown in Figure 1.

**Fig 1:**
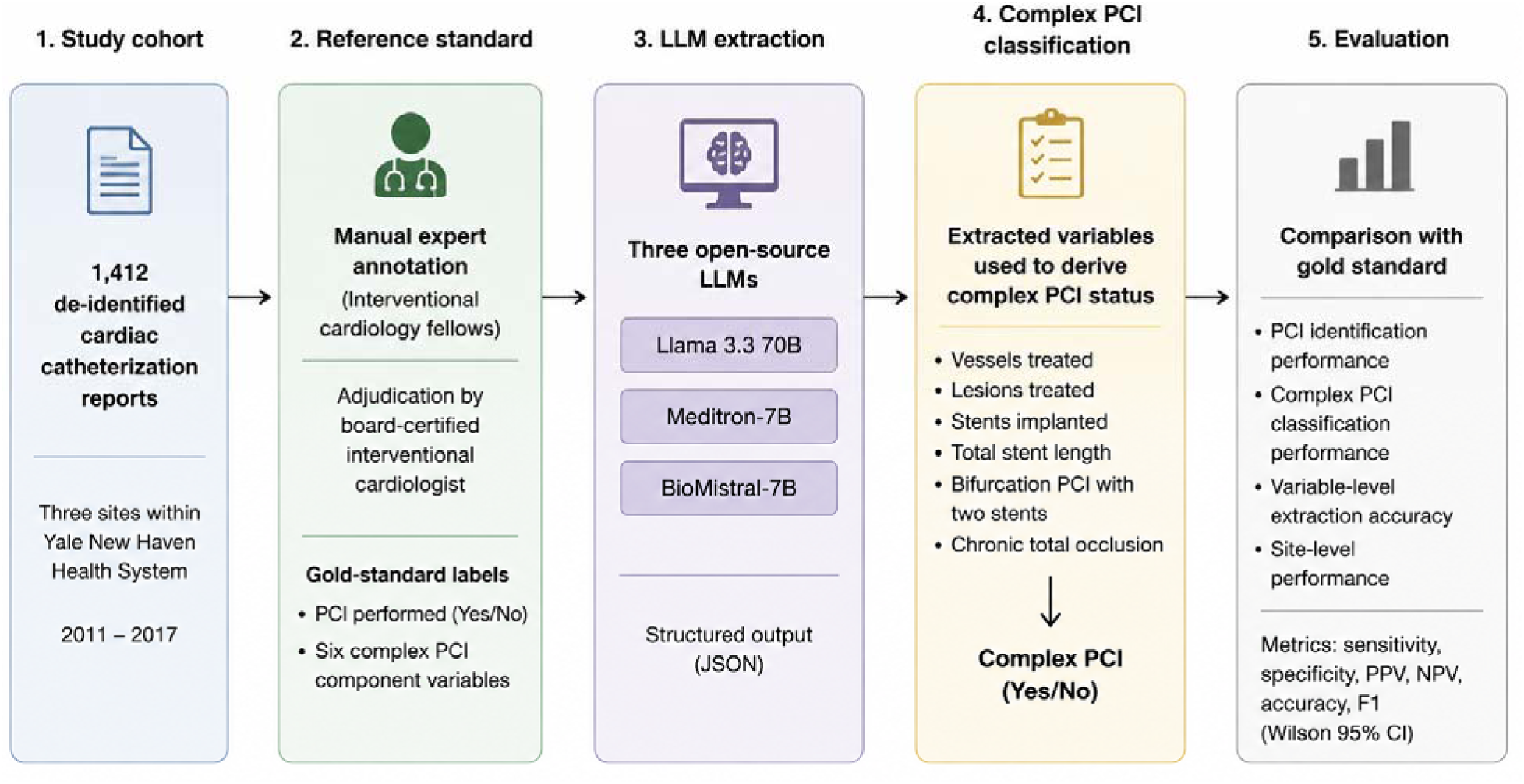
Methodology of study workflow for LLM-based extraction and evaluation of complex PCI from cath reports.

### Study Design and Dataset

The study dataset consisted of a random sample of 1412 cardiac catheterization laboratory procedure notes from three clinical sites within the Yale New Haven Health System: Yale New Haven Hospital, Lawrence and Memorial Hospital, and Bridgeport Hospital. The notes corresponded to catheterization laboratory procedures performed between 5/2011 and 1/2017 and included a range of procedure types, including diagnostic coronary angiography, peripheral vascular procedures, right heart catheterization, and percutaneous coronary interventions.

De-identified free-text reports served as the sole input text for the large language model analyses. Model evaluation included two related sequential tasks. First, models were asked to classify whether a narrative procedure note described a PCI procedure. Second, among manually verified PCI notes, models were asked to extract six variables needed to identify complex PCI based on the definition proposed by Giustino et. al.^7^ which includes number of vessels treated, number of lesions treated, number of stents implanted, bifurcation PCI with two stents implanted, total stent length, and chronic total occlusion. These variables were selected because they represent clinically meaningful procedural complexity features that may not be consistently available in simple structured fields and often require interpretation of detailed procedural text (Table 1).

**Table 1.** Variables with descriptions and examples from notes.

| Variables | Description | Examples |
| --- | --- | --- |
| PCI | Identifies whether the report describes | “Successful PCI of the mid |
| performed | a coronary PCI procedure. | LAD"; "Drug-eluting stent placed in the RCA"; "Balloon angioplasty was performed" |
| Number of vessels treated | Number of coronary vessels or major branches that underwent PCI. This captures whether the intervention was single-vessel or multivessel. | "PCI of the LAD and RCA"; "Intervention to the LCx"; "Stenting of the LAD and OM branch" |
| Number of lesions treated | Number of distinct coronary lesions treated during the procedure. This may differ from the number of vessels because more than one lesion can be treated in the same vessel. | "Proximal and mid LAD lesions were treated"; "Two RCA lesions underwent PCI"; "Stenting of the ostial LCx and OM1 lesions" |
| Number of stents implanted | Total number of coronary stents deployed during the procedure. | "One DES was deployed"; "Two overlapping stents were placed"; "A total of three stents were implanted" |
| Bifurcation with two stents implanted | Presence of bifurcation PCI treated with a two-stent strategy. | "Bifurcation PCI of the LAD/diagonal with two stents"; "Culotte stenting was performed"; "Stents placed in the main branch and side branch" |
| Total stent length | Sum of the lengths of all coronary stents implanted during the procedure, when stent dimensions were documented. | "3.0 × 38 mm and 2.75 × 24 mm DES placed"; "Overlapping 28 mm and 32 mm stents"; "Total stent length 60 mm" |
| Chronic total occlusion | Identifies whether a treated lesion was described as a chronic total occlusion or CTO, defined as a complete coronary artery blockage that is typically longstanding and requires specialized PCI techniques. | "CTO of the RCA was crossed"; "PCI of chronic total occlusion of the LAD"; "Successful CTO intervention" |

### Expert Annotation

A reference standard was created through manual review of the de-identified procedure narratives. A group of four interventional cardiology fellows (GJ, JM, PC, RT,) annotated each narrative into a predefined schema capturing PCI performed and component features of complex PCI. The annotations were then reviewed again by a board-certified interventional cardiologist (KM) to generate a highly accurate label set which was used as the reference standard for model evaluation Each report was first labeled as PCI-positive or PCI-negative. PCI-positive reports were defined as narratives documenting coronary percutaneous intervention, including balloon angioplasty, stent implantation, or other coronary interventional treatments. Diagnostic angiography alone, right heart catheterization alone, peripheral vascular intervention without coronary PCI, and other non-PCI procedures were classified as PCI-negative.

For PCI-positive reports, annotators extracted the six prespecified complex PCI variables from the narrative text. The number of vessels treated was determined from the coronary vessels or territories receiving PCI. The number of lesions treated was based on the distinct coronary lesions treated as described in the note. The number of stents implanted was calculated from documented stent deployments. Bifurcation PCI with two stents was coded as present when the report described treatment of a bifurcation lesion using a two-stent strategy. Total stent length was calculated as the sum of the lengths of all implanted coronary stents when stent dimensions were documented. Chronic total occlusion was coded as present when the treated lesion was described as a chronic total occlusion (CTO). Variables that could not be determined from the narrative were coded as Unknown.

To describe whether notes were broadly similar within and across sites, we compared narrative text using pairwise document similarity.

### Model inference

We evaluated three open-source large language models for extraction of PCI-related information from de-identified cardiac catheterization laboratory procedure narratives: a large general-purpose model - Llama 3.3 70B, and two domain-adapted biomedical language models - Meditron-7B, and BioMistral-7B. We only used open-source models to maintain patient privacy and data governance, allow reproducibility of our study results, and to enable portability of our research findings. All models were run locally on a NVIDIA A100 GPU within a secure internal computing environment. For models requiring authentication or gated access, access was obtained through the appropriate model repositories before local use. The models were used for inference only; no additional training or fine-tuning was performed.

For each de-identified narrative report, we used a standardized prompt template to guide model extraction. The prompt consisted of a brief definition of the variable, counting rules, and examples. For e.g. for PCI variable the prompt first provided a brief clinical definition of PCI, emphasizing that PCI required actual use of a therapeutic device in a coronary artery or bypass graft, such as balloon angioplasty, stent deployment, atherectomy, intravascular lithotripsy, or thrombectomy. The prompt also specified procedures that should not be counted as PCI, including diagnostic angiography alone, physiologic assessment, intravascular imaging, passage of a wire without device treatment, or attempted but unused devices.

Prompt engineering was performed before model evaluation to ensure that the instructions reflected clinical definitions and produced consistent, parsable outputs. The final template asked the model to determine whether PCI was performed and, when present, to extract prespecified complex PCI features, including number of vessels treated, number of lesions treated, number of stents implanted, bifurcation PCI with two stents, total stent length, and chronic total occlusion treatment.

Model outputs were required in a predefined JSON format using fixed field names and allowable values, with “Unknown” used when information was not available in the narrative. The same final prompt template was applied to all three models (Supplementary methods).

#### Llama 3.3 70B

Llama 3.3 70B is a large general-purpose open-weight language model. The Llama 3 model family was developed by Meta and trained on a large corpus of publicly available online text. The instruction-tuned versions of Llama 3 were further optimized for following user instructions and generating structured responses. The model was run locally using an Ollama server deployed in a Docker container on a secure internal server. For each report, the task-specific prompt was combined with the de-identified narrative report and submitted to the model for completion. Model outputs were saved for subsequent parsing and comparison with the reference standard.

#### Meditron-7B and BioMistral 7B

Meditron-7B and BioMistral 7B are medically adapted open-weight language models. These were tested to evaluate whether medical pretraining improves recognition of clinical terminology, procedural language, and abbreviations commonly present in catheterization reports.

<u>Meditron-7B</u> is based on Llama-2-7B and was further pretrained on a curated medical corpus that included selected PubMed articles, PubMed abstracts, internationally recognized medical guidelines, and general-domain text. Meditron-7B was accessed using Hugging Face Transformers after repository authentication, and the tokenizer and model weights were loaded locally on the secure server. To support inference on the available hardware, the model was run using 4-bit quantization with float16 computation. For each report, the input consisted of the task-specific prompt, the de-identified narrative report, and a structured JSON response cue. Deterministic decoding was used, with sampling disabled and temperature set to 0.0. Parsed JSON outputs were saved for evaluation.

<u>BioMistral-7B</u> is based on Mistral-7B-Instruct and was further pretrained on biomedical text from PubMed Central. BioMistral-7B was run locally using Hugging Face Transformers within the secure institutional computing environment. Like Meditron-7B, the model was loaded using 4-bit quantization with float16 computation to enable efficient local inference. The same task-specific prompts and structured input format were used. Deterministic generation was used, with sampling disabled, temperature set to 0.0, and top-p set to 1.0. Outputs were parsed to extract valid JSON when present and normalized to the predefined schema.

### Model Evaluation

Model predictions were compared with the manually adjudicated gold-standard labels. For PCI identification, reports were included when both the gold-standard and model-predicted PCI status were available. PCI performed was treated as the positive class. We calculated true-positive, false-positive, true-negative, and false-negative counts, as well as sensitivity, specificity, positive predictive value, negative predictive value, accuracy, and F1 score using standard definitions.

Extraction performance was evaluated among reports with gold-standard PCI performed. Six variables were assessed: number of vessels treated, number of lesions treated, number of stents implanted, total stent length, bifurcation PCI with two stents, and chronic total occlusion. For each variable, exact-match accuracy was calculated as the proportion of evaluable reports for which the model output matched the gold-standard value.

Each extracted variable was also converted into its corresponding binary complex PCI criterion: at least 3 vessels treated, at least 3 lesions treated, at least 3 stents implanted, total stent length greater than 60 mm, bifurcation PCI with two stents, or chronic total occlusion. For each criterion, criterion presence was treated as the positive class, and sensitivity, specificity, positive predictive value, negative predictive value, accuracy, and F1 score were calculated. These criterion-level analyses were performed overall and not stratified by clinical site.

Complex PCI status was classified as positive when at least one of the six criteria was present. A report was classified as negative only when all six criteria were available and none was present. When no criterion was positive, but one or more criteria were unavailable, complex PCI status was considered indeterminate. This rule was applied separately to the gold-standard and model-extracted variables, and complex PCI classification performance was evaluated only when both statuses could be determined.

Site-level accuracy was evaluated separately for PCI identification and complex PCI classification. Reports without an available clinical site were excluded from site-level analyses but retained in overall analyses when otherwise evaluable.

Error bars represent 95% Wilson confidence intervals for binomial proportions. Wilson intervals were calculated for sensitivity, specificity, positive predictive value, negative predictive value, accuracy, and exact-match extraction accuracy. F1 scores were reported as point estimates without confidence intervals.

Site-level heterogeneity in PCI identification was assessed using Pearson chi-square tests for equality of proportions across sites, separately for accuracy, sensitivity, and specificity. For each test, the total evaluable reports with site represented the metric-specific denominator after excluding missing-site reports: all evaluable reports for accuracy, gold-standard PCI-positive reports for sensitivity, and gold-standard PCI-negative reports for specificity. Q represents the Pearson chi-square statistic quantifying differences in performance across sites. Larger Q values indicated greater departure from equal performance across sites. An I²-style statistic was calculated as max (0, [Q − df] / Q) × 100. Values near 0% indicated little to no between-site heterogeneity and values of approximately 25%, 50% and 75% were interpreted as low, moderate and high heterogeneity respectively. NE indicated not estimable.

### Exploratory error analysis

To understand what types of ambiguous documentation were prevalent in the inaccurate classifications by the best performing model, as an exploratory analysis KM reviewed the notes of the FP and FN cases for each variable to surface possible sources of error. The sources of error from this qualitative analysis were reported as proportions.

Analysis was conducted on a secure institutional computing environment using Python. The study was approved by Yale Institutional Review Board - Protocol ID: 2000039447.

## RESULTS

### Narrative notes characteristics

The evaluation cohort included 1,412 clinical notes, of which 1,400 had an available clinical site. Notes had a median length of 718 words (IQR, 555–1,104). Note length differed by site, with shorter notes at BH and LM and longer notes at YNHH, but all sites contributed full narrative clinical documentation rather than brief structured fields.

Overall, PCI was performed in 596 of 1,412 notes (42.2%). Among PCI reports with evaluable complex PCI status, 128 of 592 (21.6%) met criteria for complex PCI. The prevalence of complex PCI was similar across sites, ranging from 21.5% to 22.2% among PCI reports. Individual complex PCI features were less common: 92 of 594 PCI reports (15.5%) involved at least 3 stents, 67 of 590 (11.4%) had total stent length greater than 60 mm, 54 of 596 (9.1%) involved up to 3 lesions, 25 of 596 (4.2%) involved chronic total occlusion, and 14 of 596 (2.3%) involved bifurcation treated with 2 stents.

Notes from the same site were more similar to one another than notes from different sites, particularly at LM, where within-site similarity was highest (median cosine similarity, 0.389; BH, 0.083; YNHH, 0.158). Across-site note similarity was lower (median, 0.051), suggesting site-level differences in documentation style. Note length also varied by site, with median word counts ranging from 495 at LM to 863 at YNHH. Despite differences in the proportion of reports involving PCI across sites, the clinical case mix among PCI reports was broadly comparable with respect to complex PCI, which accounted for 21.5% of PCI reports at BH, 22.2% at LM, and 21.6% at YNHH.

### Study cohort and evaluation structure

The final evaluation cohort included 1,412 cardiac catheterization reports from three clinical sites. By gold-standard review, 596 reports described PCI and 816 did not describe PCI. All three large language models were evaluated for identification of whether PCI had been performed. The number of evaluable reports for PCI identification was 1,386 for Llama 3 70B and 1,412 for both Meditron-7B and BioMistral-7B.

Among reports with gold-standard PCI, models were then evaluated for extraction of the six procedural variables required to classify complex PCI: number of vessels treated, number of lesions treated, number of stents implanted, total stent length, bifurcation PCI with two stents, and chronic total occlusion PCI. Because missing extracted variables could make complex PCI status indeterminate, the number of evaluable reports for complex PCI classification differed across models.

### Overall PCI identification and complex PCI classification performance

Performance differed substantially across models. For PCI identification, Llama 3 70B had the strongest overall performance, with 583 true-positive, 50 false-positive, 753 true-negative, and no false-negative classifications. This corresponded to 100.0% sensitivity, 93.8% specificity, 92.1% positive predictive value, 100.0% negative predictive value, 96.4% accuracy, and an F1 score of 95.9% (Figure 2A and Supplementary Figure 1A).

**Fig 2:**
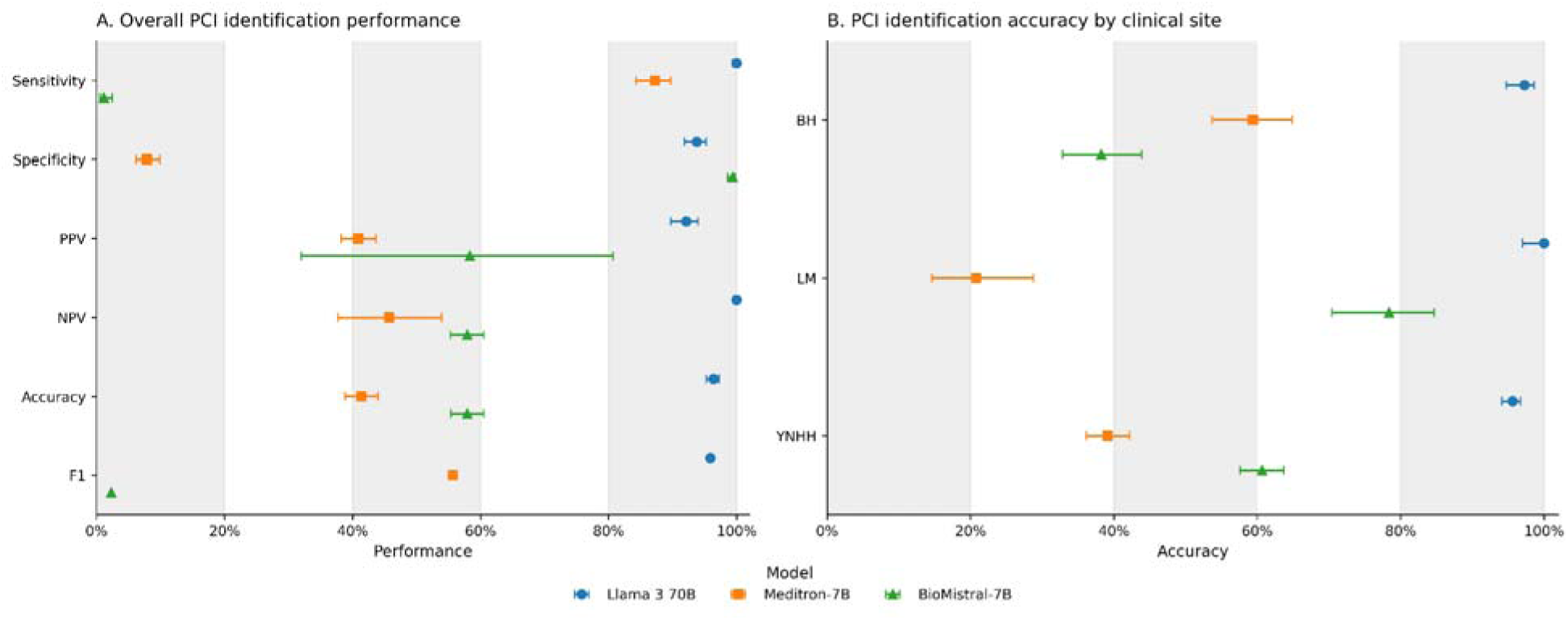
Overall and site-level performance for PCI identification. Panel A shows sensitivity, specificity, positive predictive value, negative predictive value, accuracy, and F1 score for each model. Panel B shows PCI identification accuracy by clinical site. Error bars represent 95% Wilson confidence intervals; F1 scores are displayed as point estimates without confidence intervals.

Meditron-7B identified 520 true-positive cases but produced 752 false-positive classifications. Its sensitivity was 87.2%, whereas specificity was 7.8%, positive predictive value was 40.9%, negative predictive value was 45.7%, accuracy was 41.4%, and the F1 score was 55.7%. BioMistral-7B demonstrated the opposite pattern, with 7 true-positive, 5 false-positive, 811 true-negative, and 589 false-negative classifications. Its sensitivity was 1.2%, specificity was 99.4%, positive predictive value was 58.3%, negative predictive value was 57.9%, accuracy was 57.9%, and the F1 score was 2.3%.

For complex PCI classification, Llama 3 70B again provided the best balance between sensitivity and specificity. Among 590 evaluable reports, it produced 125 true-positive, 92 false-positive, 370 true-negative, and 3 false-negative classifications. Sensitivity was 97.7%, specificity was 80.1%, positive predictive value was 57.6%, negative predictive value was 99.2%, accuracy was 83.9%, and the F1 score was 72.5% (Figure 3A and Supplementary Figure 1B).

**Fig 3:**
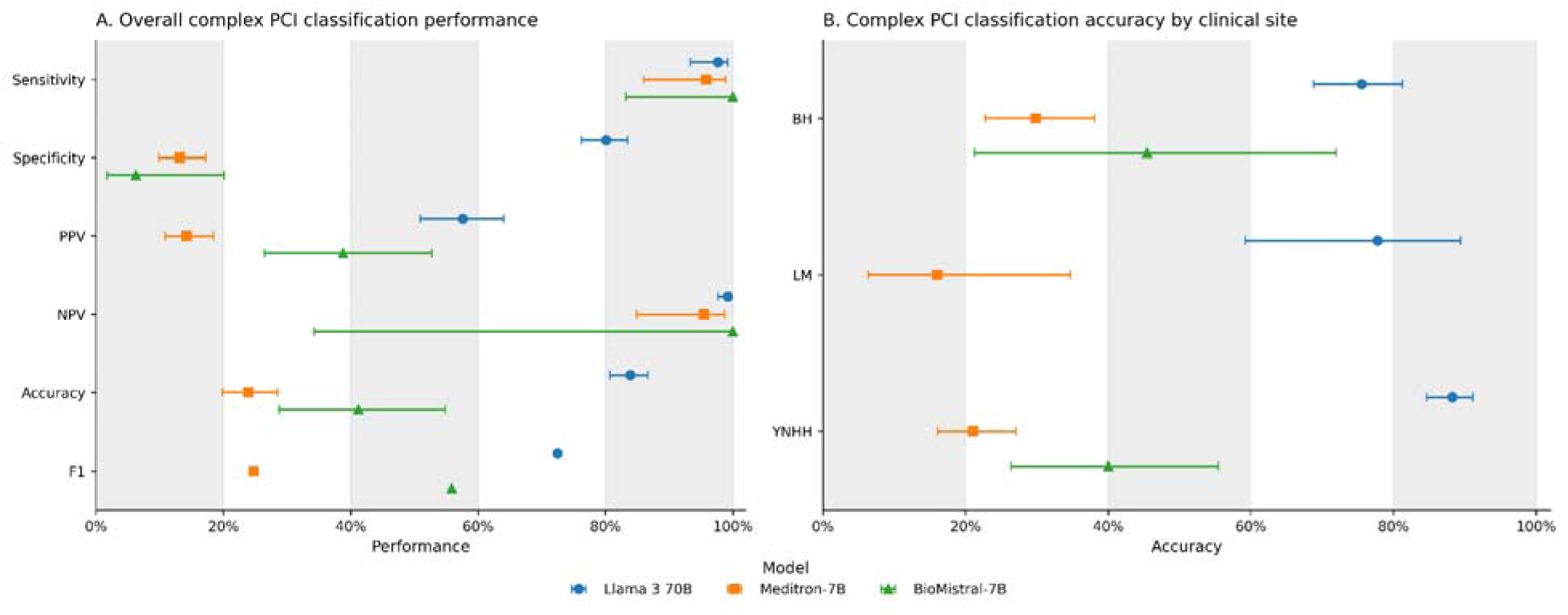
Overall and site-level performance for complex PCI classification. Panel A shows sensitivity, specificity, positive predictive value, negative predictive value, accuracy, and F1 score for each model. Panel B shows complex PCI classification accuracy by clinical site. Error bars represent 95% Wilson confidence intervals; F1 scores are displayed as point estimates without confidence intervals.

Meditron-7B had 368 evaluable reports for complex PCI classification and produced 46 true-positive, 278 false-positive, 42 true-negative, and 2 false-negative classifications. Its sensitivity was 95.8%, specificity was 13.1%, positive predictive value was 14.2%, negative predictive value was 95.5%, accuracy was 23.9%, and the F1 score was 24.7%. BioMistral-7B had only 51 evaluable reports, with 19 true-positive, 30 false-positive, 2 true-negative, and no false-negative classifications. Its sensitivity was 100.0%, specificity was 6.2%, positive predictive value was 38.8%, negative predictive value was 100.0%, accuracy was 41.2%, and the F1 score was 55.9%. The BioMistral-7B estimates should be interpreted cautiously because of the small number of evaluable reports.

### Extraction and classification of individual complex PCI criteria

Exact-match extraction accuracy varied substantially by model and variable. Llama 3 70B consistently had the highest extraction accuracy, achieving 90.1% accuracy for vessels treated, 80.7% for lesions treated, 96.8% for stents implanted, 93.8% for total stent length, 96.5% for bifurcation PCI with two stents, and 85.1% for chronic total occlusion (Figure 4).

**Fig 4:**
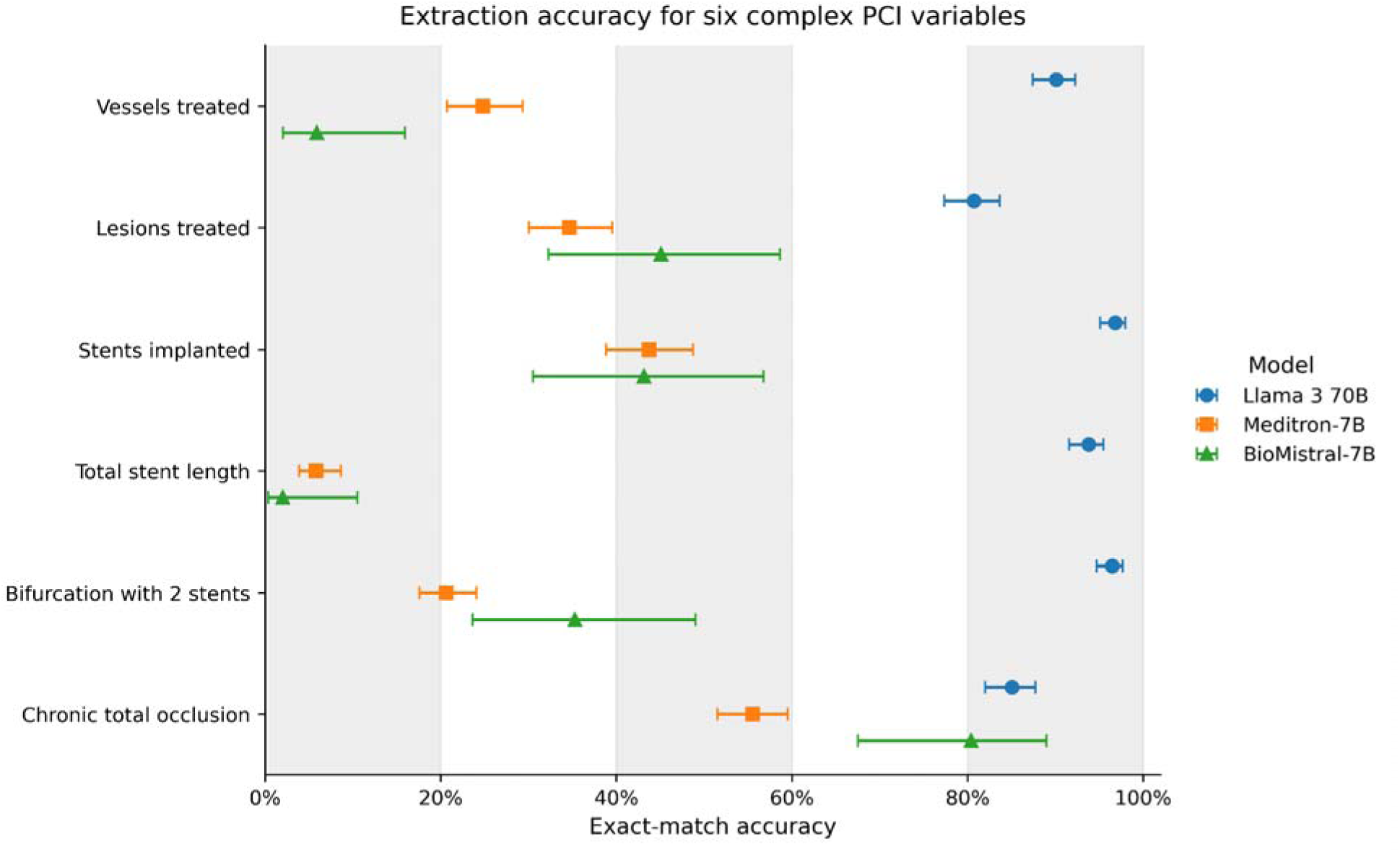
Exact-match extraction accuracy for six complex PCI variables. Exact-match accuracy is shown for number of vessels treated, number of lesions treated, number of stents implanted, total stent length, bifurcation PCI with two stents, and chronic total occlusion. Error bars represent 95% Wilson confidence intervals.

Meditron-7B had exact-match accuracies of 24.8% for vessels treated, 34.6% for lesions treated, 43.7% for stents implanted, 5.8% for total stent length, 20.6% for bifurcation PCI with two stents, and 55.5% for chronic total occlusion. Corresponding accuracies for BioMistral-7B were 5.9%, 45.1%, 43.1%, 2.0%, 35.3%, and 80.4%, respectively.

Criterion-level binary classification analyses generally showed the same model-level pattern (Supplementary Figure 2). Llama 3 70B maintained high overall accuracy across the six criteria, although positive predictive value and F1 score were lower for several uncommon criteria. Meditron-7B and BioMistral-7B frequently demonstrated high sensitivity or high apparent accuracy for selected criteria but low specificity or positive predictive value. This pattern was particularly evident for rare complex PCI features, for which a model could achieve high accuracy by predominantly classifying reports as criterion negative. Full criterion-level metrics and confusion-matrix counts are presented in Supplementary Table 1.

### Performance by clinical site

For PCI identification, Llama 3 70B maintained high accuracy across all three clinical sites, with approximate accuracies of 97.1% at BH, 100.0% at LM, and 95.5% at YNHH (Figure 2B). Meditron-7B demonstrated substantially greater variation, with approximate accuracies of 59.6%, 20.9%, and 39.3% at BH, LM, and YNHH, respectively. BioMistral-7B also varied across sites, with approximate accuracies of 38.2% at BH, 78.3% at LM, and 60.7% at YNHH.

Similarly, Llama 3 70B had the highest accuracy at each site for complex PCI classification (Figure 3B), with approximate accuracies of 75.6% at BH, 77.7% at LM, and 88.2% at YNHH. Meditron-7B achieved approximate accuracies of 29.8%, 16.0%, and 21.1%, respectively. BioMistral-7B achieved approximate accuracies of 45.4% at BH and 40.0% at YNHH; an estimate was not available at LM because no reports had evaluable gold-standard and model-derived complex PCI classifications.

Reports with missing clinical sites were excluded from the site-level analyses but retained in the corresponding overall analyses when otherwise evaluable.

### Site-level performance and heterogeneity

After excluding reports with missing clinical sites, PCI identification performance varied significantly across clinical sites for all three models, with the greatest heterogeneity observed for BioMistral-7B and Meditron-7B. This variation was driven primarily by differences in accuracy and specificity rather than sensitivity. The counts of reports per site also play a significant role in the underlying heterogeneity.

For complex PCI classification, significant site-level heterogeneity was observed for Llama 3 70B, particularly for accuracy and specificity, whereas heterogeneity was not statistically significant for Meditron-7B or BioMistral-7B (Table 2).

**Table 2:** Site-level heterogeneity in PCI identification performance and complex PCI classification.

| Model | Metric | Total | Total | Q statistic | P value | I2-style |
| --- | --- | --- | --- | --- | --- | --- |

|  |  | evaluabl<br>e sites | evaluable<br>reports<br>with site |  |  | statistic<br>, % |
| --- | --- | --- | --- | --- | --- | --- |
| **PCI identification performance** |  |  |  |  |  |  |
| Llama 3 70B | Accuracy | 3 | 1374 | 6.96 | 0.031 | 71.3 |
|  | Sensitivity | 3 | 583 | NE | NE | NE |
|  | Specificity | 3 | 791 | 7.55 | 0.023 | 73.5 |
| Meditron-7B | Accuracy | 3 | 1400 | 62.87 | 0 | 96.8 |
|  | Sensitivity | 3 | 596 | 3.24 | 0.198 | 38.2 |
|  | Specificity | 3 | 804 | 10.2 | 0.006 | 80.4 |
| BioMistral-7B | Accuracy | 3 | 1400 | 71.01 | 0 | 97.2 |
|  | Sensitivity | 3 | 596 | 1.4 | 0.495 | 0 |
|  | Specificity | 3 | 804 | 1.77 | 0.413 | 0 |
| **Complex PCI classification** |  |  |  |  |  |  |
| Llama 3 70B | Accuracy | 3 | 590 | 15.39 | <0.001 | 87.01 |
|  | Sensitivity | 3 | 128 | 1.66 | 0.435 | 0 |
|  | Specificity | 3 | 462 | 18.84 | <0.001 | 89.38 |
| Meditron-7B | Accuracy | 3 | 368 | 4.396 | 0.111 | 54.51 |
|  | Sensitivity | 3 | 48 | 5.175 | 0.075 | 61.35 |
|  | Specificity | 3 | 320 | 4.769 | 0.092 | 58.06 |
| BioMistral-7B | Accuracy | 2 | 51 | 0.106 | 0.745 | 0 |
|  | Sensitivity | 2 | 19 | NE | NE | NE |
|  | Specificity | 2 | 32 | 0.492 | 0.483 | 0 |

### Exploratory error analysis

Potential reasons for misclassification identified on manual review for PCI identification and the 6 complex PCI variables are shown in Table 3.

**Table 3:** Manual error analysis of LLM predictions.

| **Variable** | **Potential reasons for misclassification identified on manual review** |
| --- | --- |
| Identifying PCI | IVUS or FFR only procedure misclassified as PCI (70%)<br>Non-coronary vascular interventions (18%)<br>Misinterpretation of historical or referential mentions of PCI (10%)<br>Unknown (2%) |
| Number of vessels treated | 2 lesions in 1 vessel territory counted as 2 vessels (39%)<br>Unknown (25%)<br>Non-PCI vessel FFR/IVUS counted (22%)<br>Ambiguous documentation (3%)<br>Multiple (3%)<br>LM bifurcation vessel count misinterpreted (3%)<br>Misinterpretation of historical or referential mentions of additional vessel PCI (2%)<br>Attempted PCI in additional vessel (2%) |
| Number of lesions treated* | Unknown (81%)<br>Ambiguous documentation (12%)<br>Non-PCI vessel FFR/IVUS counted (7%) |
| Number of stents implanted | Ambiguous documentation (37%)<br>Balloon brand name with length mentioned but not referred to as balloon (16%)<br>Unknown (16%)<br>One of the stents was attempted but not deployed (11%)<br>Prior stent lengths reported (11%)<br>Zero stents implanted, prediction output as unknown (11%) |
| Total stent length | Ambiguous documentation (41%)<br>Unknown (38%)<br>Balloon brand name with length mentioned but not referred to as balloon (11%)<br>One of the stents was attempted but not deployed (5%)<br>Prior stent lengths reported (5%) |
| Bifurcation with two stents implanted | Unknown (95%) – use of words like ‘bifurcation lesion’, several instances of stent in one vessel and balloon angioplasty of branch<br>Ambiguous documentation (5%) |
| Chronic total occlusion (CTO) | Unknown (51%)<br>Total occlusion in the setting of myocardial infarction (22%)<br>Non-PCI CTO vessel present on angiography (22%)<br>Mention of a prior CTO procedure (3%)<br>Ambiguous description of CTO (3%) |

\*Only cases where number of vessels was accurately predicted but number of lesions was inaccurate were reviewed

## DISCUSSION

In this study, we demonstrate that a contemporary open-source large language model, Llama 3.3 70B, can accurately extract complex percutaneous coronary intervention (PCI) variables from unstructured cardiac catheterization reports. Llama 3.3 70B demonstrated vastly better performance than smaller domain-specific models - Meditron-7B and BioMistral-7B. These findings add to a rapidly evolving body of literature suggesting that large, general-purpose language models can perform structured clinical data extraction tasks with high fidelity, even in specialized domains such as interventional cardiology.

Prior work in clinical natural language processing largely relied on rule-based systems or traditional deep learning approaches (e.g., BERT-based models), which require extensive annotation and are often require task-specific annotation and model development and may have limited portability across documentation environments.^8^ Recent studies have shown that large language models can outperform these approaches in some clinical information extraction tasks, particularly when training data are limited or when tasks involve nuanced contextual interpretation, particularly in heterogeneous clinical corpora.^5^ For example, LLM-based systems have demonstrated good accuracy in extracting structured variables from a variety of medical text such as operative notes^9,10^, pathology reports^4,11^, radiology reports^12,13^, and clinical trial reports.^14^ The application of such systems to PCI notes remains comparatively limited. One prior study showed promising performance of LLMs for identifying PCI procedural variables, however, the number of annotated PCI notes were smaller limiting performance assessment, and importantly the LLMs used were not open-source.^6^ Our study included a much larger sample, used open-source models which are critical for health-care applications, and performed the computations in a secure HIPAA-compliant environment.

Our findings extend the literature in several important ways. First, we focus on complex PCI variables, which represent a particularly challenging extraction task due to variability in terminology (e.g., bifurcation strategies, lesion characteristics), implicit documentation, and dependence on procedural context. The strong performance observed with a large model suggests that LLMs can capture higher-order clinical reasoning embedded in free-text narratives, rather than simply identifying surface-level entities. Second, the marked performance gap between the larger general-purpose model and the smaller medical-domain models suggests that model capacity and general language-reasoning ability may may, for some complex extraction tasks, outweigh the benefits of domain-specific pretraining alone.^15^ This aligns with emerging evidence that larger models demonstrate superior generalization and contextual understanding, even in specialized medical applications.

The observed variation in performance across complex PCI variables likely reflects both differences in documentation characteristics and the underlying cognitive demands placed on large language models. Variables with the highest accuracy—such as number of stents, and stent length—are typically explicitly stated, numerically defined, and locally documented within discrete portions of the procedure note, making them well-suited to direct extraction. These fields require relatively limited contextual integration and align closely with the strengths of LLMs in pattern recognition and span-level retrieval. In contrast, variables with lower accuracy, such as chronic total occlusion and lesions treated, often require interpretation of more heterogeneous language, integration of information across multiple sections of the note, and application of clinical definitions that may not be explicitly labeled.^3,6,16^ For example, identifying a chronic total occlusion may depend on implicit descriptors (e.g., “100% occluded,” but differentiating it from an acute setting, or historical context), while lesion counting can be sensitive to how operators describe contiguous versus discrete diseased segments, overlap across vessels, or procedural intent. These tasks place greater demands on multi-step reasoning, coreference resolution, and consistent application of domain-specific rules, increasing susceptibility to error. Our error analysis revealed that most misclassifications stemmed from the interaction between heterogeneous or fragmented procedural documentation and the need for contextual interpretation beyond simple entity extraction. Broadly, our findings highlight that LLM performance remains highest for well-structured, explicitly documented variables, and declines for latent constructs that must be inferred, underscoring the potential value of hybrid approaches that combine LLM-based extraction with rule-based logic or post-processing constraints for more complex PCI phenotyping.

These findings have important implications for the future of cardiovascular research and quality measurement. Manual abstraction of PCI variables from cath reports is time-intensive, costly, and limits scalability of registries and observational studies.^1^ LLM-based extraction offers the potential to enable scalable automated phenotyping from routine clinical documentation, facilitating more granular and timely evaluation of procedural complexity and outcomes. Future work should focus on (1) development of hybrid approaches combining rule-based constraints with LLM reasoning^17^, (2) integration with structured data pipelines and registry workflows, (3) external validation across diverse health systems, and (4) evaluation of downstream impact on clinical research and quality reporting. In addition, emerging strategies such as model ensembles^18^ or retrieval-based approaches may further improve performance for extraction tasks requiring integration of dispersed or complex contextual information, although benefits appear task-dependent.^19,20^

Despite these promising results, several limitations merit consideration. LLM-based extraction is inherently probabilistic and may produce occasional errors, particularly for infrequently documented or ambiguously described variables. Prior work has demonstrated that even small inaccuracies in extracted variables can propagate and meaningfully impact downstream analyses^21,22^, especially when variables are used for risk modeling or comparative effectiveness research. Additionally, performance may degrade as document length and information density increase, particularly when relevant information is dispersed across the note or when the task requires identifying missing information^23^, and variability in documentation practices across institutions may affect generalizability.^24^ Importantly, while our study demonstrates strong performance in a retrospective setting, prospective validation and human-in-the-loop workflows^25^ remain essential before clinical or research deployment.

Our study has limitations. Our study was limited to one health system; however, we compared performance across 3 sites with distinct note structures increasing the external validity of our results. We performed prompt engineering with a few notes for rapid prototyping, and prompt engineering at scale could lead to further improvements in accuracy. However, the focus of our study was a high-level feasibility assessment of using LLMs for extracting complex procedural variables. Development of an EHR-ready LLM based pipeline to extract variables at scale will need to incorporate detailed prompt engineering, fine tuning, experimenting with hybrid pipelines combining LLM extraction with rules-based systems, and human-in-the-loop integration. We limited our testing to 3 open-source models as these could be run in a secure environment. Other LLMs could have better performance.

## CONCLUSION

Large language models—particularly high-capacity models—show substantial promise for automated extraction of complex interventional cardiology variables from unstructured text. While important challenges remain, these tools have the potential to fundamentally transform clinical data abstraction and enable more scalable, high-resolution cardiovascular research.

## Datasets/Code availability statement

Due to privacy and IRB constraints, raw data is not publicly distributed. Link to our code is available at https://github.com/itsjustnilay/complex-pci-llm-evaluation/.

## Funding Sources

Dr Murugiah received support from the National Heart, Lung, and Blood Institute of the National Institutes of Health (under award K08HL157727).

## Conflict of Interest Disclosure

Dr Krumholz, in the past two years, has received options for OpenEvidence, Element Science and Identifeye and payments from F-Prime for advisory roles. He was a co-founder of and held equity in Hugo Health. He is a co-founder of and holds equity in Refactor Health and ENSIGHT-AI. He is a co-founder of medRxiv and is on the Board of openRxiv (non-paid, volunteer). He is associated with research contracts through Yale University from Janssen, Kenvue, Novartis, and Pfizer.

## Data Availability

The data are PHI and cannot be shared. The prompts used and the code used to conduct the project will be shared on GitHub

## Notes

### Author Declarations

Yale University Institutional Review Board

### Summary of Updates

Additional figures. Changes to supplement.

