## Supplementary materials for "Automated Identification of Complex Percutaneous Coronary Intervention from Cardiac Catheterization Reports using Large Language Models"

**Contents:**

- **Supplementary Methods**
- **Supplementary Table 1**
- **Supplementary Figure 1**
- **Supplementary Figure 2**

**Abbreviations:** CTO, chronic total occlusion; FN, false negative; FP, false positive; NPV, negative predictive value; PCI, percutaneous coronary intervention; PPV, positive predictive value; TN, true negative; TP, true positive.

**Supplementary Methods**

**Supplementary Methods 1. Model output schema**

{
 "PCI_performed": "Yes|No",
 "vessels_treated": "0..N|Unknown",
 "lesions_treated": "0..N|Unknown",
 "stents_implanted": "0..N|Unknown",
 "bifurcation_with_2_stents": "Yes|No|Unknown",
 "total_stent_length_mm": "number|Unknown",
 "chronic_total_occlusion": "Yes|No|Unknown",
 "complex_pci": "Yes|No|Unknown"
}

**Supplementary Methods 2. Prompt for PCI identification**

You are analyzing narrative reports describing heart procedures. Your task is to determine whether a Percutaneous Coronary Intervention (PCI) was performed.

Definition of PCI: A PCI is a percutaneous procedure to treat a stenosis or blockage in a coronary artery or coronary bypass graft using a device to mechanically restore or improve blood flow. PCI requires that a device was actually used in the coronary artery or graft (not just attempted to be passed into the artery or graft).

Specifically:

- Balloon = only if the balloon was used to dilate a stenosis in an artery or graft

- Stent = only if deployed in an artery or graft

- Atherectomy, intravascular lithotripsy, brachytherapy = only if activated in an artery or graft

- Thrombectomy = only if the device was passed into an artery or graft and aspiration/suction performed

- Do NOT count if a device was only attempted but never used

- Do NOT count diagnostic procedures alone (FFR, iFR, pressure wire, or intravascular imaging alone) as PCI

- Do NOT count passing a wire across the stenosis alone without use of a device as PCI

Output:

Return only a JSON object, with no explanation. Always include the key exactly as shown. Do not add extra keys.

Format:

{

"PCI_performed": "Yes" or "No"

}

Here is the narrative report:

**Supplementary Methods 3. Prompt for extraction of complex PCI variables**

You are given narrative reports of PCI procedures. Your task is to:

1. Extract six required clinical details.

2. Use those details to determine whether the case qualifies as Complex PCI (see criteria).

3. Output only a JSON object without additional explanations. Always include all keys, no blanks. Use "Unknown" if unclear.

Step 1: Extract the six required details

1. vessels_treated (integer or "Unknown")

- Count the number of unique major coronary arteries or bypass grafts treated with PCI (e.g., LM, LAD, LCX, RCA, LIMA to LAD, SVG-OM etc.).

- A stent to the main vessel such as the LAD and a diagonal branch counts as 1 vessel not 2. Similarly a stent in OM1 and OM2 count as 1 (LCX system)

- Examples:

- "PCI to LAD and RCA" → 2

- "Treated proximal and mid LCX" → 1

- "Stented SVG-OM ostium"	→ 1

- "Stented from LAD into diagonal" → 1

2. lesions_treated (integer or "Unknown")

- Count distinct lesion sites treated (even within the same vessel system). If one lesion extends continuously from one vessel into another (e.g., LM into LCX ostium), it is considered a single contiguous lesion

- Examples:

- "Proximal and mid LAD lesions treated" → 2

- "Three lesions across LAD and RCA" → 3

- "Lesion extending from distal LM into LCX" → 1

- "Lesion in ostial LM and a lesion in proximal LCX treated" → 2

3. stents_implanted (integer or "Unknown")

- Count all stents deployed during the entire procedure.

- Examples:

- "A single 3.0 x 28 mm stent was deployed" → 1

- "Two overlapping drug-eluting stents were placed" → 2

4. bifurcation_with_2_stents ("Yes" or "No")

- Identify if a bifurcation (branch point) was treated using a two-stent technique.

- Look for terms like: DK crush, culotte, mini-crush, T-stenting and look if 2 or more stents were deployed in the bifurcation.

- Examples:

- "Culotte technique for LAD/D1 bifurcation" → Yes

- "Provisional stenting" → No

5. total_stent_length_mm (integer or "Unknown")

- Sum the lengths (in mm) of all stents deployed in all the arteries.

- Examples:

- "3.0 x 18 mm and 2.5 x 24 mm stents" → 42

- "Total stent length was 78 mm" → 78

6. chronic_total_occlusion ("Yes" or "No")

- Identify if a chronic total occlusion (CTO) was treated.

- CTO = complete blockage >3 months

- Look for:

- "CTO of RCA treated"

- "Chronic total occlusion intervention performed"

Step 2: Determine if this was a Complex PCI

A PCI is considered complex if ANY ONE of the following criteria are met:

- 3 or more vessels treated

- 3 or more lesions treated

- 3 or more stents implanted

- Bifurcation treated with 2 stents

- Total stent length > 60 mm

- CTO treated

Step 3: Output a JSON object in the below format

Output Format:

{

"vessels_treated": ...,

"lesions_treated": ...,

"stents_implanted": ...,

"bifurcation_with_2_stents": "...",

"total_stent_length_mm": ...,

"chronic_total_occlusion": "...",

"complex_pci": "Yes" or "No"

}

**Supplementary Table 1**

| **Model** | **Evaluable reports** | **TP** | **FP** | **TN** | **FN** | **Sensitivity%** | **Specificity%** | **PPV%** | **NPV%** | **Accuracy%** | **F1 %** |
| --- | --- | --- | --- | --- | --- | --- | --- | --- | --- | --- | --- |
| **PCI Identification** | | | | | | | | | | | |
| Llama 3 70B | 1386 | 583 | 50 | 753 | 0 | 100 | 93.8 | 92.1 | 100 | 96.4 | 95.9 |
| Meditron-7B | 1412 | 520 | 752 | 64 | 76 | 87.2 | 7.8 | 40.9 | 45.7 | 41.4 | 55.7 |
| BioMistral-7B | 1412 | 7 | 5 | 811 | 589 | 1.2 | 99.4 | 58.3 | 57.9 | 57.9 | 2.3 |
| **Complex PCI classification** | | | | | | | | | | | |
| Llama 3 70B | 590 | 125 | 92 | 370 | 3 | 97.7 | 80.1 | 57.6 | 99.2 | 83.9 | 72.5 |
| Meditron-7B | 368 | 46 | 278 | 42 | 2 | 95.8 | 13.1 | 14.2 | 95.5 | 23.9 | 24.7 |
| BioMistral-7B | 51 | 19 | 30 | 2 | 0 | 100 | 6.2 | 38.8 | 100 | 41.2 | 55.9 |
| **Criterion: Vessels treated >=3** | | | | | | | | | | | |
| Llama 3 70B | 596 | 8 | 26 | 562 | 0 | 100 | 95.6 | 23.5 | 100 | 95.6 | 38.1 |
| Meditron-7B | 383 | 2 | 153 | 228 | 0 | 100 | 59.8 | 1.3 | 100 | 60.1 | 2.5 |
| BioMistral-7B | 51 | 0 | 48 | 3 | 0 | 0 | 5.9 | 0 | 100 | 5.9 | 0 |
| **Criterion: Lesions treated >=3** | | | | | | | | | | | |
| Llama 3 70B | 596 | 51 | 51 | 491 | 3 | 94.4 | 90.6 | 50 | 99.4 | 90.9 | 65.4 |
| Meditron-7B | 384 | 14 | 152 | 215 | 3 | 82.4 | 58.6 | 8.4 | 98.6 | 59.6 | 15.3 |
| BioMistral-7B | 51 | 4 | 25 | 21 | 1 | 80 | 45.7 | 13.8 | 95.5 | 49 | 23.5 |
| **Criterion: Stents implanted >=3** | | | | | | | | | | | |
| Llama 3 70B | 592 | 91 | 3 | 497 | 1 | 98.9 | 99.4 | 96.8 | 99.8 | 99.3 | 97.8 |
| Meditron-7B | 382 | 29 | 138 | 212 | 3 | 90.6 | 60.6 | 17.4 | 98.6 | 63.1 | 29.1 |
| BioMistral-7B | 51 | 11 | 21 | 19 | 0 | 100 | 47.5 | 34.4 | 100 | 58.8 | 51.2 |
| **Criterion: Total stent length >60 mm** | | | | | | | | | | | |
| Llama 3 70B | 587 | 67 | 5 | 515 | 0 | 100 | 99 | 93.1 | 100 | 99.1 | 96.4 |
| Meditron-7B | 380 | 2 | 1 | 356 | 21 | 8.7 | 99.7 | 66.7 | 94.4 | 94.2 | 15.4 |
| BioMistral-7B | 50 | 5 | 7 | 29 | 9 | 35.7 | 80.6 | 41.7 | 76.3 | 68 | 38.5 |
| **Criterion: Bifurcation with 2 stents** | | | | | | | | | | | |
| Llama 3 70B | 596 | 13 | 20 | 562 | 1 | 92.9 | 96.6 | 39.4 | 99.8 | 96.5 | 55.3 |
| Meditron-7B | 368 | 6 | 245 | 117 | 0 | 100 | 32.3 | 2.4 | 100 | 33.4 | 4.7 |
| BioMistral-7B | 51 | 1 | 33 | 17 | 0 | 100 | 34 | 2.9 | 100 | 35.3 | 5.7 |
| **Criterion: Chronic total occlusion** | | | | | | | | | | | |
| Llama 3 70B | 596 | 19 | 95 | 476 | 6 | 76 | 83.4 | 16.7 | 98.8 | 83.1 | 27.3 |
| Meditron-7B | 368 | 3 | 29 | 328 | 8 | 27.3 | 91.9 | 9.4 | 97.6 | 89.9 | 14 |
| BioMistral-7B | 51 | 1 | 9 | 40 | 1 | 50 | 81.6 | 10 | 97.6 | 80.4 | 16.7 |

**Supplementary Table 1**. Model performance metrics for PCI identification, overall complex PCI classification, and classification of each individual complex PCI criterion. Reported measures include the number of evaluable reports, true-positive, false-positive, true-negative, and false-negative counts, sensitivity, specificity, positive predictive value, negative predictive value, accuracy, and F1 score. For PCI identification, PCI performed was treated as the positive class. For complex PCI classification, meeting at least one complex PCI criterion was treated as the positive class. For criterion-level analyses, presence of the corresponding criterion was treated as the positive class. Analyses of complex PCI and its component criteria were restricted to reports with gold-standard PCI performed. Overall analyses retained reports without an available clinical site when otherwise evaluable.

**Supplementary Figure 1**

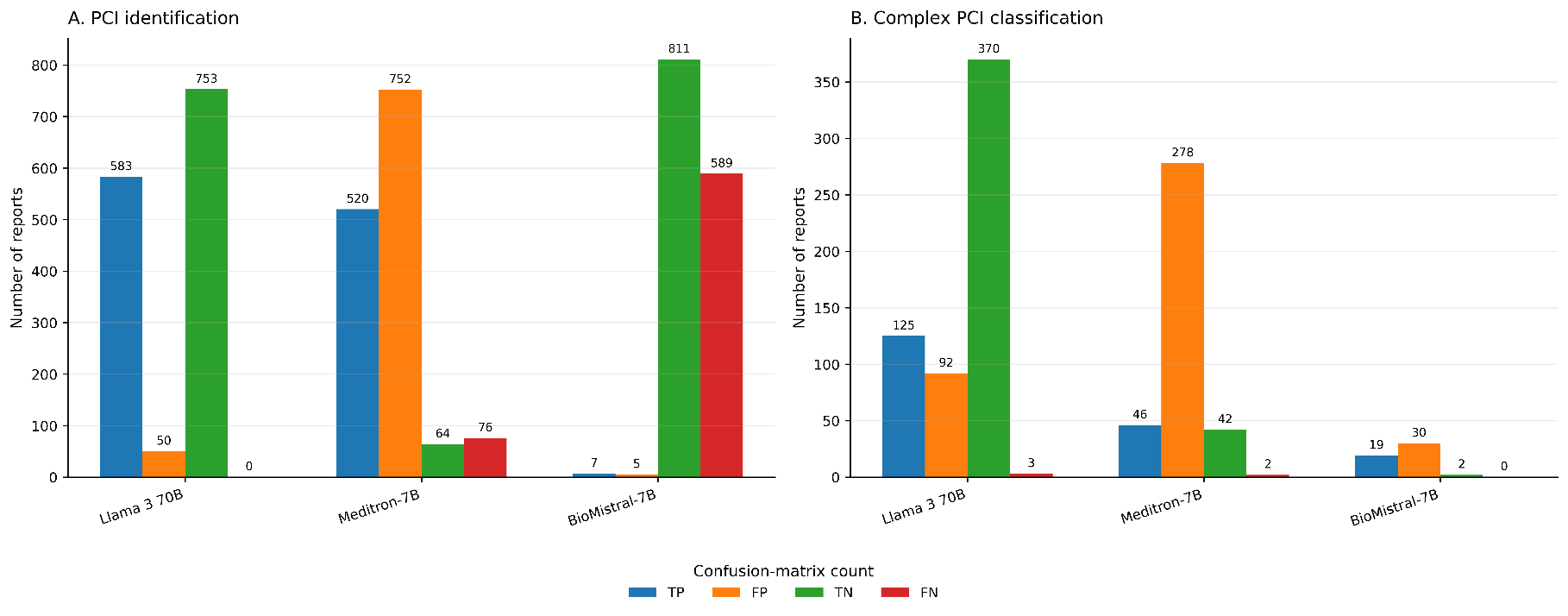

**Supplementary Figure 1. Confusion-matrix counts for PCI identification and complex PCI classification.** Panel A shows true-positive, false-positive, true-negative, and false-negative counts for identification of whether PCI was performed. Panel B shows the corresponding confusion-matrix counts for complex PCI classification among reports with gold-standard PCI performed and evaluable gold-standard and model-derived complex PCI status. Counts are shown separately for Llama 3 70B, Meditron-7B, and BioMistral-7B. Differences in total evaluable counts reflect missing or indeterminate model outputs.

**Supplementary Figure 2**
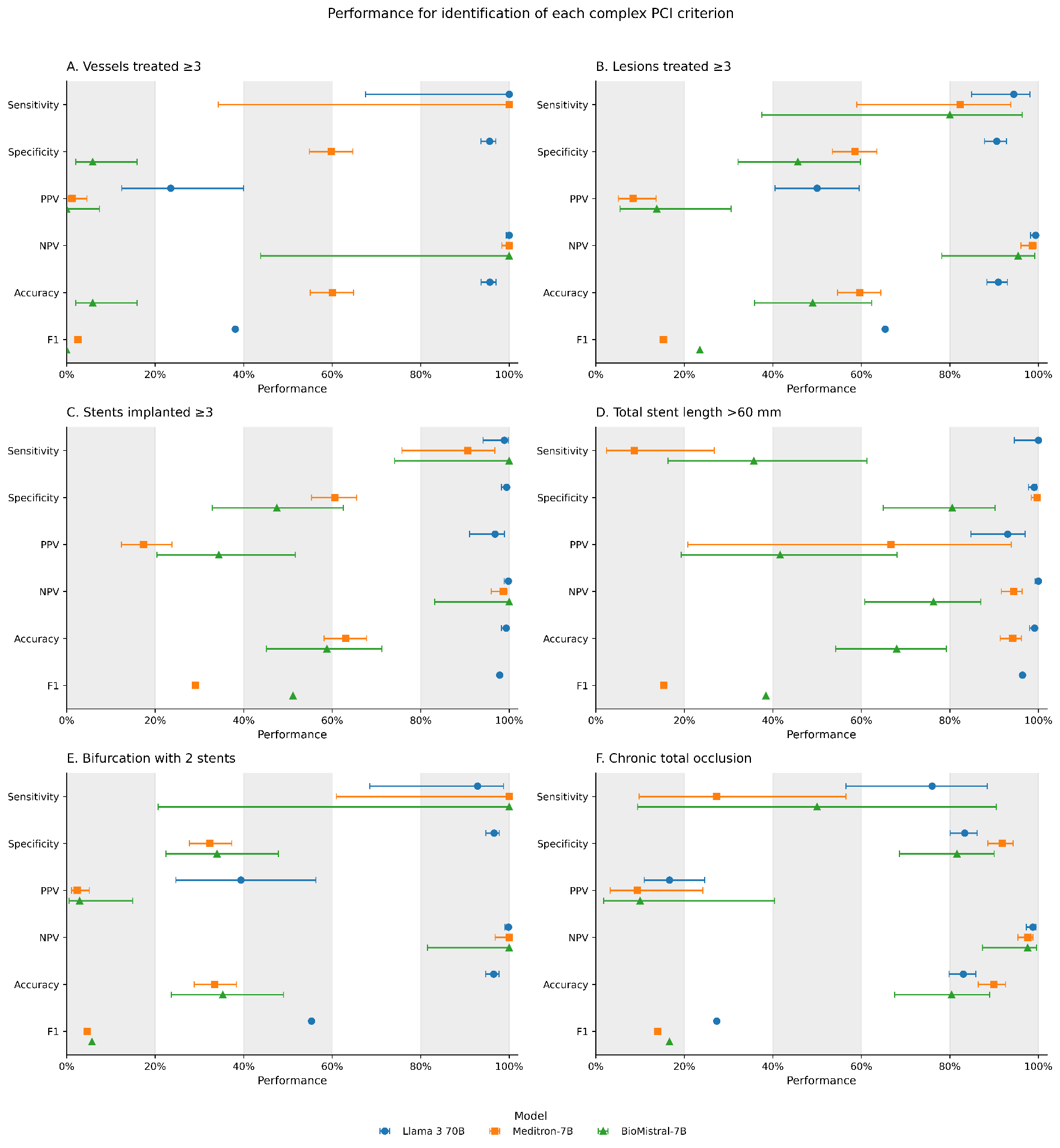

**Supplementary Figure 2. Performance for classification of each individual complex PCI criterion.** Forest plots show sensitivity, specificity, positive predictive value, negative predictive value, accuracy, and F1 score for classification of each of the six complex PCI criteria: Panel A, at least 3 vessels treated; B, at least 3 lesions treated; C, at least 3 stents implanted; D, total stent length greater than 60 mm; Panel E, bifurcation PCI treated with 2 stents; and Panel F, chronic total occlusion. Criterion presence was treated as the positive class. Analyses were restricted to reports with gold-standard PCI performed and evaluable gold-standard and model-predicted values for the corresponding criterion. Error bars represent 95% Wilson confidence intervals. F1 scores are shown as point estimates without confidence intervals.
